# Large-scale population analysis of SARS-CoV-2 whole genome sequences reveals host-mediated viral evolution with emergence of mutations in the viral Spike protein associated with elevated mortality rates

**DOI:** 10.1101/2020.10.23.20218511

**Authors:** Carlos Farkas, Andy Mella, Jody J. Haigh

## Abstract

**Background:** We aimed to further characterize and analyze in depth intra-host variation and founder variants of SARS-CoV-2 worldwide up until August 2020, by examining in excess of 94,000 SARS-CoV-2 viral sequences in order to understand SARS-CoV-2 variant evolution, how these variants arose and identify any increased mortality associated with these variants.

**Methods and Findings:** We combined worldwide sequencing data from GISAID and Sequence Read Archive (SRA) repositories and discovered SARS-CoV-2 hypermutation occurring in less than 2% of COVID19 patients, likely caused by host mechanisms involved APOBEC3G complexes and intra-host microdiversity. Most of this intra-host variation occurring in SARS-CoV-2 are predicted to change viral proteins with defined variant signatures, demonstrating that SARS-CoV-2 can be actively shaped by the host immune system to varying degrees. At the global population level, several SARS-CoV-2 proteins such as Nsp2, 3C-like proteinase, ORF3a and ORF8 are under active evolution, as evidenced by their increased πN/ πS ratios per geographical region. Importantly, two emergent variants: V1176F in co-occurrence with D614G mutation in the viral Spike protein, and S477N, located in the Receptor Binding Domain (RBD) of the Spike protein, are associated with high fatality rates and are increasingly spreading throughout the world. The S477N variant arose quickly in Australia and experimental data support that this variant increases Spike protein fitness and its binding to ACE2.

**Conclusions:** SARS-CoV-2 is evolving non-randomly, and human hosts shape emergent variants with positive fitness that can easily spread into the population. We propose that V1776F and S477N variants occurring in the Spike protein are two novel mutations occurring in SARS-CoV-2 and may pose significant public health concerns in the future.

**Author Summary:** We have developed an efficient bioinformatics pipeline that has allowed us obtain the most complete picture to date of how the SARS-CoV-2 virus has changed during the last eight month global pandemic and will continue to change in the near future. We characterized the importance of the host immune response in shaping viral variants at different degrees, evidenced by hypermutation responses on SARS-CoV-2 in less than 2% of infections and positive selection of several viral proteins by geographical region. We underscore how human hosts are shaping emergent variants with positive fitness that can easily spread into the population, evidenced by variants V1176F and S477N, located in the stalk and receptor binding domains of the Spike protein, respectively. Variant V1176 is associated with increased mortality rates in Brazil and variant S477N is associated with increased mortality rates over the world. In addition, it has been experimentally demonstrated that S477N variant increase fitness of Spike protein and its binding with ACE2, thus predicting to increase virulence of SARS-CoV-2. This limits the concept of ‘herd immunity’ proposals and re-emphasize the need to limit the spread of the virus to avoid emergence of more virulent forms of SARS-CoV-2 that can spread worldwide.

## Introduction

The novel SARS-CoV-2 coronavirus that causes COVID19 has surpassed 34 million infections worldwide within nine months of pandemic, resulting in more than one million deaths until September 2020 (https://coronavirus.jhu.edu/map.html) (1). In-depth characterization of this virus is urgently needed to improve outbreak surveillance, vaccine development and for effective treatments now and in the immediate future. SARS-CoV-2 is a positive single-stranded RNA virus (+ssRNA) with a crown-like appearance observed by electron microscopy that is due the presence of the of spike glycoproteins on the lipid bilayer envelope (2, 3). Another three transmembrane proteins are incorporated into the envelope: small envelope protein (E), matrix protein (M), and nucleocapsid protein (N) (4). As seen with SARS-CoV-1, SARS-CoV-2 binds through its Spike glycoprotein to cell membrane-bound angiotensin-converting enzyme 2 (ACE2) for entry into host cells (5–8). Advancements in COVID19 treatments have been recently developed including Remdesivir, a nucleoside analog that inhibits viral RNA-dependent RNA polymerase and is an effective treatment to reduce viral titers in rhesus macaques that is clinically approved for COVID19 treatment (9). As well, either wild-type or catalytically inactive ACE2 has been demonstrated to block viral entry *in vitro*, and are proposed as promising treatments (10, 11). A remaining question is how the human humoral immune response develops after SARS-CoV-2 infection. Studies in Iceland have shown that around 90% of infected patients develop antiviral antibodies that last up to four months (12), but it has also been suggested that around one third of the seropositive infections are asymptomatic and become antibody-negative early in the convalescence period (13). Also, the unexpectedly low secondary infection risk reported for SARS-CoV-2 infection suggests innate immune responses are active in humans (14, 15). To explore host– SARS-CoV-2 interactions at the genetic level it is useful to analyze viral sequencing results per individual and at the population level. Initiatives such as GISAID (https://www.gisaid.org/) (16, 17) and the Sequence Read Archive (SRA, https://www.ncbi.nlm.nih.gov/sra) have been storing SARS-CoV-2 sequencing datasets worldwide from the beginning of the pandemic starting in January 2020, allowing researchers to track fixed variants and follow viral evolution by geographical region. The unprecedented amount of SARS-CoV-2 whole genome sequencing data can help to 1) characterize viral variants that occur within a given host, 2) understand variant fixation in a given population and 3) understand how the virus changes over time. In fact, the Spike protein mutation D614G global transition that occurred very recently was discovered in this way and is associated with higher viral titers and higher fatality rates (18, 19). Thus, it is probable that more mutations are to be discovered by tracking SARS-CoV-2 genomic changes globally. In this study we aimed to characterize in depth intra-host variation and population-fixed variants worldwide up until the beginning of August 2020 by using over 76,000 SARS-CoV-2 sequences and 17,500 sequencing datasets from GISAID and SRA repositories, respectively. First, we found evidence for SARS-CoV-2 hypermutation, occurring in less than 2% of COVID19 patients. This mechanism is predicted to inactivate the virus and is likely caused by host mechanisms involved APOBEC3G complexes and intra-host microdiversity, where G>T transversions and C>T transitions are frequent signatures observed both in hypermutant and non-hypermutant samples. These results suggest that SARS-CoV-2 is actively shaped by the host immune system to varying degrees. From a population context, several SARS-CoV-2 proteins such as Nsp2, 3C-like proteinase, ORF3a and ORF8 are under active evolution, evidenced by their increasing πN/ πS ratios.

Noteworthy, most of the population-fixed variants in SARS-CoV-2 are predicted to destabilize viral proteins, as already reported for other RNA viruses. Of these variants, those occurring in the ORF3a (Q57H), Nucleocapsid (I292T, RG203KR) and Spike protein (V1176F) have a positive association with increased mortality ratios in populations from Saudi-Arabia and Brazil, respectively. In particular, the V1176F variant co-occurs with the D614G mutation in the Spike protein mutation in Brazil and arose independently in at least in three independent SARS-CoV-2 clades. This variant is predicted to stabilize the SARS-CoV-2 Spike trimmer complex and confer flexibility to the stalk domain of the trimmer, potentially facilitating Spike binding properties to ACE2. Also, this variant is associated with increased mortality ratios in Brazil and is increasingly spreading throughout the world. Similarly, the emerging variant S477N occurring in the Receptor Binding Domain, dramatically increase its frequency and became dominant in Australia within two months. Experimental data support that S477N increase both fitness and binding to ACE2 receptor, explaining its selection among other viruses in Australia. S477N also is presently spreading across countries and is associated with higher fatalities throughout the world. We propose that these variants are novel mutations occurring in SARS-CoV-2 and their spread may pose serious concerns in public health in the future of the pandemic.

## Methods

### Data and Code Availability

76,553 FASTA genomes and associated sequencing metadata were downloaded from GISAID database from January 1, 2019 until August 3, 2020, specifying “human” as source host (https://www.gisaid.org/). The associated sequencing metadata including major variants per sample are available at Supplementary Table 1. 974 Brazilian FASTA sequences were downloaded from GISAID database from January 1, 2019 until September 25, 2020, specifying “human” as source host and “South America / Brazil” as location. Acknowledgements to all laboratories/consortia involved in the generation of GISAID genomes used in this study are listed in Supplementary Table 2. 17,560 sequencing datasets were downloaded from Sequence Read Archive Repository (SRA, https://www.ncbi.nlm.nih.gov/sars-cov-2/) From December 1, 2019 until July 28, 2020. Associated sequencing run accessions, sequencing metadata and related BioProjects are listed in Supplementary Table 3. The code generated during this study to replicate most of the computational calculations performed in this manuscript is available at the following github repository: https://github.com/cfarkas/SARS-CoV-2-freebayes.

### Next-generation sequencing and FASTA dataset processing

To process next generation sequencing datasets, we employed our pipeline (SARS-CoV-2_freebayes) consisting in a bash/UNIX script that pipes several programs in sequential order. Imputed list of SRA accessions is processed with sra-tools, https://github.com/ncbi/sra-tools), generating compressed FASTQ files per sequencing, automatically trimmed with fastp tool (20). Minimap2 splice-aware aligner in preset mode -ax sr (21) align each trimmed fastq file against a provided reference genome (Wuhan-Hu-1, GenBank Accession: MN908947.3). The resulting BAM files were sorted and indexed by using Samtools (22). Freebayes, as frequency-based pooled caller (-F 0.49) (https://github.com/ekg/freebayes) (23) perform variant calling on every sorted BAM file, obtaining major frequency viral variants per genome in VCF format,. Then, the Jacquard program (https://jacquard.readthedocs.io/en/v0.42/index.html) in python environment (24) merges every VCF file containing variants associated to each bam file into a single VCF file, containing aggregated variants from all genomes. Viral frequencies were recalculated in the merge VCF file by using several UNIX tools (25), in combination with vcflib (https://github.com/vcflib/vcflib). Variants per genome reported in the resulting file “logfile_variants_SRA_freebayes” were used to construct Figure 1B using GraphPad Prism 8 software (https://www.graphpad.com/scientific-software/prism/). GISAID FASTA genomes were processed in a similar manner. We preprocess a single GISAID genome collection with SeqKit (26) to decompose a single FASTA file into individual FASTA files, each file containing a single genome. Then, Minimap2 aligner with preset -ax asm5 (21) align every FASTA genome against SARS-CoV-2 reference genome. Freebayes variant caller with --min-alternate-count 1 (C 1) option (https://github.com/ekg/freebayes) perform variant calling on each BAM file, outputting variants in VCF format. With these operations, major frequency viral variants in VCF format are obtained from each FASTA genome. Then, variants are aggregated into a single VCF file, as described with Jacquard. Figure 1A graph was constructed by using variants per genome, reported in the output file “logfile_variants_GISAID_freebayes”, inputted into the GraphPad Prism 8 software. All these computational analyses are described here: https://github.com/cfarkas/SARS-CoV-2-freebayes (case examples I and II, respectively).

**Figure 1:**
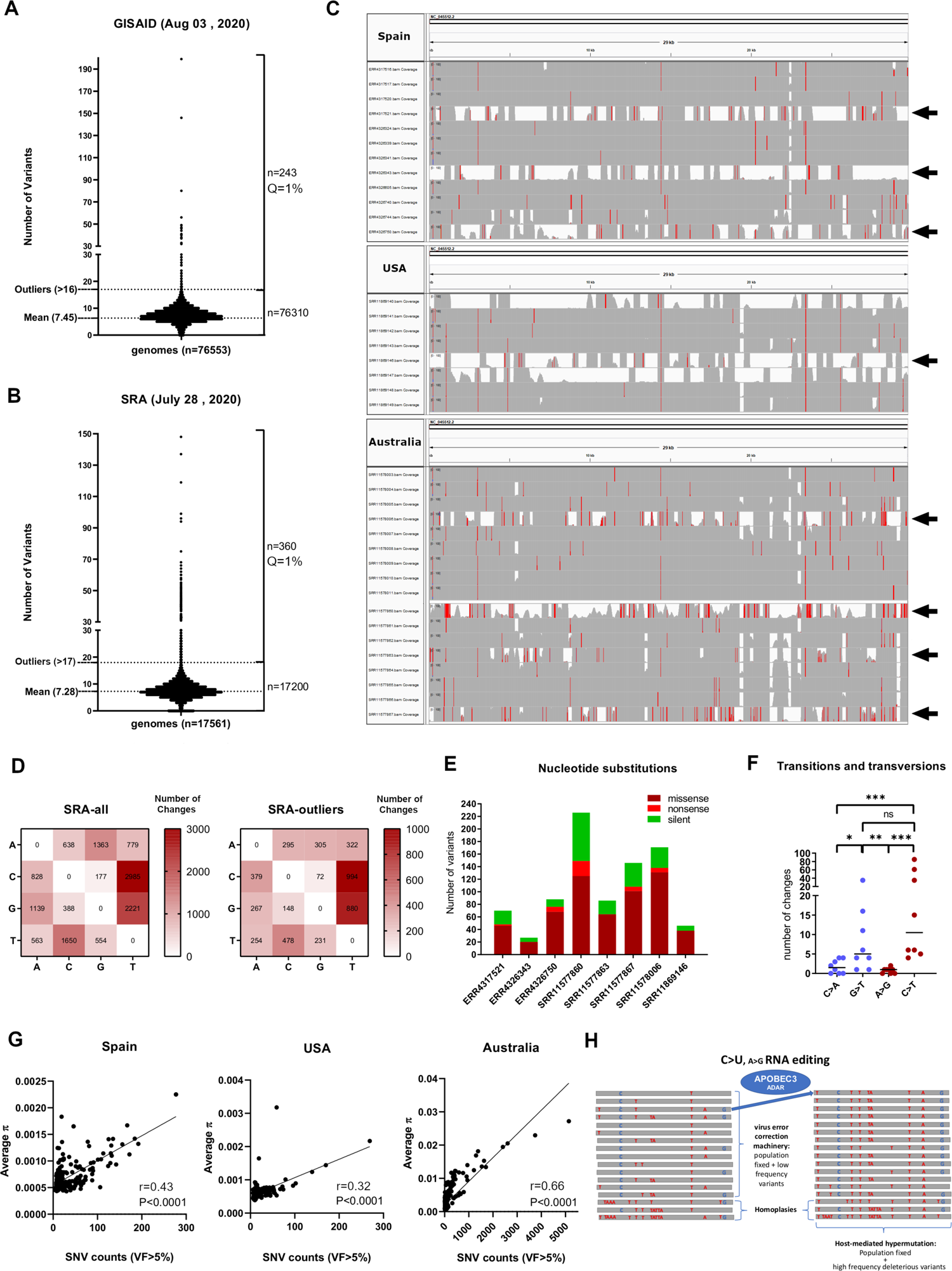
Intra-host variation in SARS-CoV-2 genomes worldwide reveal microdiversity and hypermutation likely elicited by APOBEC3G complexes. A) Mayor viral frequency variants (via a consensus calling approach) for 75,563 SARS-CoV-2 GISAID genomes, separated by non-outliers (n=76,310) and outliers (n=243, Q=1%, Grubbs’s test). Mean and outlier number of variants are depicted at left. B) Same as A for next generation sequencing (NGS) datasets downloaded from SRA (n=17,500). C) IGV snapshots of outliers and non-outlier NGS samples from B. Outliers samples are depicted with black arrows, exceeding number of variants from non-outliers. Single nucleotide polymorphisms are depicted in red if nucleotide differs from the reference sequence in greater than 50% of quality weighted reads. D) Nucleotide change frequencies from 17,560 SRA NGS aggregated variants (left) and from 360 aggregated outlier variants (right), both annotated with SnpEff program. Frequency boxes are colored from white to dark red as number of changes increases. E) Nucleotide substitution frequencies in each of the eight outlier samples indicated with black arrows in C) grouped by silent (green), missense (dark red) and nonsense (red). F) Transitions and transversions occurring in the latter samples, denoted with blue and red, respectively. Significance of comparisons were assessed with Mann-Whitney test (P<0.05 *, P<0.01 **, P<0.001 ***, P>0.05 ns). G) Correlation between Average nucleotide diversity (π) provided by inStrain program and SNV counts (VF>5%) for Spain (n=374, left), USA (n=215, middle) and Australian NGS samples (n=397, right). In the three countries, the two variables tend to increase together (see r values). The significance thresholds were the following: P<0.05 *, P<0.01 **, P<0.001 ***, P<0.0001 ****, P>0.05 ns. H) Proposed model of how APOBEC3G/ADAR complex can lead to hypermutation of SARS-CoV-2 (C>U and A>G editing). The RNA editing can be accompanied by intra-host diversity (low frequency variants) and homoplasy (different viral lineages emerged after the infection), maintained at low frequency due the virus error correction machinery.

### Variant Visualization

The Integrative Genomics Viewer (IGV) software (http://software.broadinstitute.org/software/igv/home) was used to visualize next generation sequencing alignments in bam format (27–29). To visualize mayor viral frequency variants, the variant frequency threshold was set at 0.49.

### SnpEff annotation

Merged variants from GISAID genomes (n=76563) were annotated by using in a repurposed version of SnpEff program, available in the Galaxy server (30–32). The resulting annotated VCF file was parsed by using conventional UNIX tools. All these computational analyses are described here: https://github.com/cfarkas/SARS-CoV-2-freebayes (case example III).

### πN/ πS calculation

We estimated nonsynonymous and synonymous nucleotide diversity (πN and πS, respectively) in 1279, 6841, 46042, 17989, 1205 and 2924 GISAID FASTA genomes from Africa, Asia, Europe, North America, South America, and Oceania, respectively. The FASTA files were processed from the alignment to the variant calling step as described in “GISAID FASTA dataset processing section”. All these computational analyses are described here: https://github.com/cfarkas/SARS-CoV-2-freebayes (case example IV).

### Microdiversity and low frequency viral variants

We estimated nucleotide diversity in 397, 448 and 308 next generation sequencing (NGS) samples from Australia, Spain, and USA populations, respectively by using aligned reads per sample in BAM format against SARS-CoV-2 reference genome. These BAM files were inputted in loop to InStrain program (https://instrain.readthedocs.io/en/latest/) (33), obtaining several parameters such as analysis of coverage, microdiversity, SNV linkage, and sensitive SNP detection, among others. As recommended by inStrain, we analyzed only sequencing samples with sufficient breadth of coverage (>0.9), resulting in 397, 374 and 216 NGS samples from Australia, Spain and from USA, respectively. The list of the NGS samples in the three populations, including the referred calculations are detailed in the spreadsheet inStrain_results.xlsx, available here: https://github.com/cfarkas/SARS-CoV-2-freebayes. We correlated in each country the number of variants with viral frequency > 5% against the nucleotide diversity (π) by using Spearman correlation. Spearman’s correlation coefficients (r) and confident p-values (P, to discard random sampling) were calculated in GraphPad Prism 8. The significance thresholds were as follows: P<0.05 *, P<0.01 **, P<0.001 ***, P<0.0001 ****, P>0.05 ns. All these computational analyses are described here: https://github.com/cfarkas/SARS-CoV-2-freebayes (case example V).

### SNP-mortality associations

We downloaded 7634 genomes with associated metadata from GISAID until September 28, 2020 and we grouped the genomes from released/deceased patients per country (India, Saudi Arabia, USA, and Brazil, respectively). Then, we parsed genomes and associated metadata by country (in particular, deceased and released cases) by using a combination of standard UNIX tools, vcflib (https://github.com/vcflib/vcflib) and BEDOPS (34). After these steps, we uploaded to the Galaxy server (https://usegalaxy.org/) the resulting output per country (Deceased-Released.subset file) (31, 35) and we performed Fisher’s exact test to identified variants with a significant difference in the viral frequencies between the groups (snpFreq program, https://rdrr.io/github/lvclark/SNPfreq/). P values from Fisher’s exact test were converted with to negative logarithm in base 10 by using R version 3.6.3 (https://www.r-project.org/). The latter values per variant were used to construct graph from Figures 3E and 3H, respectively by using GraphPad Prism 8 software. All computational steps required for these analyses are available here: https://github.com/cfarkas/SARS-CoV-2-freebayes (case example VI).

### Phylogenetic Tree Construction

We downloaded 393 SAR2-CoV-2 GISAID genomes containing variant V1176F until September 25, 2020 from GISAID database (https://www.gisaid.org/). We filtered countries with at least two sequences per country, leading 358 sequences encompassing five country/regions (Brazil, Scottland, USA, Australia, and Gibraltar, respectively). MAFFT multiple sequence alignment program version 7.271 (36, 37) was used to align FASTA sequences against Wuhan-Wu-1 reference genome (GenBank Accession: MN908947.3) by using the --auto --thread −1 -- keeplength –addfragments flags. Fasttree version 2.1 (38) was used to infer an approximately-maximum-likelihood phylogenetic tree from the aligned sequences in fasta format, by using heuristic neighbor-joining clustering method (38) and the Jukes-Cantor model of evolution (39). Visualization and editing of the phylogenetic tree was performed by using Interactive Tree of Life server (iTOL), collapsing all clades whose average branch length distance was below 0.0002 (40, 41).

### Free energy estimation calculations

SARS-CoV-2 protein models for nsp2, nsp3, nsp4, 3C-Like proteinase, nsp6, nsp7, nsp8, RNA-dependent RNA polymerase, Helicase, Spike protein trimmer, ORF3a, ORF6, ORF8 and Nucleocapsid, respectively were accessed and downloaded from I-TASSER server (https://zhanglab.ccmb.med.umich.edu/COVID-19/) on June 20, 2020, in Protein Data Bank (PDB) format. These models were generated by the C-I-TASSER pipeline (42–45). We calculated the free energy of Gibbs upon variant changes (ΔΔG; ΔG_wild-type_ – ΔG_variant_, in kcal/mol) from all missense variants listed in Figure 3A using SARS-CoV-2 protein structures as inputs for the Foldx5 program (46, 47). We repaired every PDB by using the following command: foldx --command=RepairPDB --pdb=name-of-protein.pdb --ionStrength=0.05 -- pH=7 --water=CRYSTAL --vdwDesign=2 --out-pdb=true --pdbHydrogens=false. Then, we modelled the variant and calculated the free energy upon aminoacid changes as follows: foldx -- command=BuildModel --pdb=name-of-protein.pdb --mutant-file=individual_list.txt -- ionStrength=0.05 --pH=7 --water=CRYSTAL --vdwDesign=2 --out-pdb=true -- pdbHydrogens=false --numberOfRuns=30, where individual_list.txt contain the aminoacid change (as example, for a serine/asparagine change occurring in the three chains of the spike protein trimmer: SA477N,SB477N,SC477N;). We classified the energetic effects as follows (in kcal/mol): highly stabilizing (< −1.84), stabilizing (−1.84 to −0.92), slightly stabilizing (−0.92 to −0.46), neutral (−0.46 to +0.46), slightly destabilizing (+0.46 to +0.92), destabilizing (+0.92 to +1.84), and highly destabilizing (> +1.84) (48). Additionally, the effect of D614G/V1176F variants on protein–protein interaction energy in the full Spike protein trimmer and the effect of S477N variant in the Receptor Binding Domain (RBD) complexed with ACE2 dimer were assessed by submitting the referred structures on mCSM-PPI2 server (http://biosig.unimelb.edu.au/mcsm_ppi2/) (49).

### Molecular dynamics simulations

We computed molecular dynamics simulations of the wild-type (Valine at position 1176) and 1176F (Phenylalanine at position 1176) stalk domain trimmers from the spike protein (aminoacids 1130-1273). The full Spike protein trimmer was obtained from I-TASSER and the variant V1176F was modelled by using Foldx5, as previously described in the Free energy estimation calculations section (--command=BuildModel, first outputted model). The wild-type and F1176 variant trimmers were subjected to molecular dynamics by using GROMACS/2020.3 version, in gpu mode (http://manual.gromacs.org/documentation/) (50, 51) in the supercomputer infrastructure LEFTRARU NLHPC (ECM-02), allocating one node with total 44 cores (logical) and one compatible GPU (NVIDIA Tesla V100-PCIE-16GB). The trajectories were visualized by using VMD 1.9.3 (52). As example, for a given pdb (molecula_1.pdb), the commands used to perform the molecular dynamics are the following:

srun -p general gmx pdb2gmx -f molecula_1.pdb -o molecula_2.gro -water spce srun -p general gmx editconf -f molecula_2.gro -o molecula_3.gro -c -d 1.0 -bt cubic

srun -p general gmx solvate -cp molecula_3.gro -cs spc216.gro -o molecula_4.gro -p topol.top srun -p general gmx grompp -f ions.mdp -c molecula_4.gro -p topol.top -o ions.tpr

srun -p general gmx genion -s ions.tpr -o molecula_5.gro -p topol.top -pname NA -nname CL - neutral

gmx grompp -f 1.mdp -c molecula_5.gro -p topol.top -o em.tpr gmx mdrun -nt 20 -nb gpu -deffnm em #EM

gmx grompp -f 2.mdp -c em.gro -r em.gro -p topol.top -o nvt.tpr gmx mdrun -nt 20 -nb gpu -deffnm nvt # NPT

gmx grompp -f 3.mdp -c nvt.gro -r nvt.gro -t nvt.cpt -p topol.top -o npt.tpr

gmx mdrun -nt 20 -nb gpu -deffnm npt # NVT

gmx grompp -f 4.mdp -c npt.gro -t npt.cpt -p topol.top -o md_0_1.tpr gmx mdrun -nt 20 -nb gpu -deffnm md_0_1 # MD

Where commands starting with “srun” were executed directly in the cluster and the remaining steps were submitted via SLURM workload manager (https://slurm.schedmd.com/documentation.html). We choose forcefield OPLS-AA/L all-atom force field (2001 aminoacid dihedrals) for step one, and SOLVENT for step five (choice 13, SOL). Atom clashed in the system were minimized by the steepest descent method (53), until potential energy were below 1000 kJ/(mol*nm). We considered a cutoff of 1.0 nm for non-LJ bonded interactions under periodic boundary conditions (PBC). NVT ensemble (constant Number of particles, Volume, and Temperature) was performed setting no pressure coupling and modified Berendsen thermostat at 300K, respectively. The NPT ensemble was used to keep the constant pressure at 1 bar, using the Parrinello-Rahman barostat and temperature at 300 K, using the modified Berendsen thermostat, respectively. Long-range electrostatic forces were considered using the Particle Mesh Ewald for long-range electrostatics method (54). Both equilibrations were performed for 5000 picoseconds (5 nanoseconds). The total energy, temperature, pressure and the of the stalk domain trimmers were used to corroborate both system equilibrations. After these steps, production dynamics were carried out for 20 nanoseconds, by using the leap-frog algorithm with an integration step of 2 femtoseconds, as motion setting. Bonds were fixed using the P-LINCS method, with constrained H-bonds (55, 56). Root mean square deviation (RMSD) and radius of gyration (Rg) were obtained with the following commands, respectively:

gmx rms -s md.tpr -f md_0_1.xtc -o rmsd.xvg -tu ns # Choose twice 4 (“Backbone”) gmx gyrate -s md.tpr -f md_0_1.xtc -o gyrate.xvg # Group 1 (Protein)

The xvg file records per picosecond were used to plot graphs from Figure 3C, on GraphPad Prism 8 software. PDB, solvated molecules (.gro) and correspondent compressed gromacs trajectories (with or without periodic border conditions) are available here: https://usegalaxy.org/u/carlosfarkas/h/sars-cov-2-proteins-and-trayectories.

### Protein Visualization and Rendering

The Spike protein trimmer image related to Figure 3C was rendered online with the EzMol server (http://www.sbg.bio.ic.ac.uk/ezmol/) (57).

### Statistical analysis

All statistical analyses were carried out by using GraphPad Prism 8 software (https://www.graphpad.com/scientific-software/prism/). Mann-Whitney test was employed to test data following non normal distribution. The significance thresholds were the following: P<0.05 *, P<0.01 **, P<0.001 ***, P>0.05 ns. We interpreted Spearman nonparametric correlations analyses as follows: perfect correlation (1), the two variables tend to increase or decrease together (0 to 1), The two variables do not vary together at all (0), One variable increases as the other decreases (−1 to 0), and perfect inverse correlations (−1). We computed an approximate P value because in all correlations we employed more than 17 pair values. The significance thresholds were the following: P<0.05 *, P<0.01 **, P<0.001 ***, P<0.0001 ****, P>0.05 ns. We employed robust regression and outlier removal (ROUT) method (58) to remove outliers from stacks of data, with a strict false discovery ratio (Q=1%). SNP-mortality correlations were assessed by using the snpFreq program (https://rdrr.io/github/lvclark/SNPfreq/), implemented in the R language (https://www.r-project.org/). We employed the Fisher’s exact test to identified variants with a significant difference in the viral frequencies between deceased-released groups. We used as a significance threshold a false discovery rate (q-value) of 0.005.

## Results

### Worldwide Intra-host variation in SARS-CoV-2 genomes reveal microdiversity and hypermutation likely elicited by APOBEC3G/ADAR complexes

To trace intra-host viral variation worldwide, we downloaded and analyzed 76,553 SARS-CoV-2 genome sequences available in the GISAID database up until August 3, 2020 (**Supplementary Table 1**, see Acknowledgements in **Supplementary Table 2**). We also downloaded and analyzed 17,560 next-generation sequencing datasets from Sequence Read Archive (SRA) available until July 28, 2020 (**Supplementary Table 3**). SARS-CoV-2 genomes from GISAID accounted for the presence of major viral frequency variants (via a consensus calling approach) and the next-generation sequencing datasets (NGS) also allowed us to analyze intra-host microdiversity given the depth of sequencing. By analyzing the occurrence of major viral alleles per SARS-CoV-2 genome, both sources consistently demonstrate on average 7-8 viral variants with major alleles per genome (viral frequency > 0.5) (see “mean” in **Figure 1A** and **1B**, for SRA and GISAID datasets, respectively), demonstrating that our variant calling pipeline is reliable to call major viral alleles from FASTA and NGS datasets, respectively (see https://github.com/cfarkas/SARS-CoV-2-freebayes). The distribution from both sources also identified outliers with more than 16 viral sequence variants per genome including some samples harboring more than 100 variants per genome, greatly surpassing the average (2% and 0.3% in SRA and GIDAID sequencing datasets, see “outliers” in **Figure 1A and 1B**, respectively, Q=1% Grubbs’s test). Integrative genomics viewer (IGV) snapshots of outlier samples from Spain, USA and Australian sequencing datasets clearly show hypermutability to varying degrees (see samples with black arrows, **Figure 1C**). Australian outlier samples represent an extreme case of hypermutability (see **Figure 1C**, bottom). 16,307 aggregated variants from SRA datasets reflect most recurrent single nucleotide substitutions occurring in all genomes from SRA repository are enriched by the C>U (C>T) transitions and G>T (G>U) transversions, changes already reported for SARS-CoV-2 and MERS-CoV genomes (59) and is likely elicited by APOBEC deaminases, as already reported (60). (**Figure 1D**, left). The latter observation is also consistent for genomes containing outlier number of variants from SRA (**Figure 1D**, right). Most of the nucleotide substitutions harboring outlier samples from Figure 1C correspond to missense/nonsense variants rather than silent variants (**Figure 1E**) and are enriched in C>T (C>U) changes as well, consistent with latter observations (**Figure 1F**). Also, G>T (G>U) transversions are significantly present, implying a different mechanism than APOBEC3G editing and likely exerted by ADAR deaminase (see discussion). We estimated intra-host nucleotide diversity occurring in 397, 374 and 215 next generation sequencing samples from Australia, Spain and USA populations, respectively by using aligned reads per sample against the SARS-CoV-2 reference genome (Wuhan-Hu-1, GenBank Accession: MN908947.3). This calculation has been already validated to capture intra-host viral microdiversity, overcoming sequencing errors (61). In the three populations, average nucleotide diversity positively correlates with the number of Single Nucleotide Variants (SNVs) with viral frequencies over 5% (Spearman correlation, r values from 0.32-0.66, P<0.0001). The latter supports the existence of intra-host minor variants and therefore SARS-CoV-2 quasi-species, coexisting within the same host (62–65) (**Figure 1G**). We hypothesized the existence of hypermutants can be explained by APOBEC3G/ADAR-mediating RNA editing at early stages of SARS-CoV-2 infection (C>U and A>G editing) accompanied by intra-host diversity, i.e. low frequency variance and homoplasy (different viral lineages emerged after the infection) that probably are maintained at low frequency due the virus error correction machinery (**Figure 1H**). Overall, we propose that human hosts substantially contribute to shape SARS-CoV-2 genetic diversity. Although microdiversity is probably one of the main sources of SARS-CoV-2 evolution, this is accompanied by RNA-editing at different levels, with SARS-CoV-2 RNA hypermutation as an extreme case of the latter.

### Non-neutral codon changes actively shape evolution of several SARS-CoV-2 proteins

We next analyzed all intra-host major viral alleles occurring in SARS-CoV-2 genomes worldwide, by using the GISAID consensus called variants. 23,269 aggregated variants from GISAID demonstrate that overall, C>U and A>G edition are the predominant nucleotide changes (**Figure 2A**), also present in GISAID samples with outlier number of variants per genome (**Figure 2B**). These changes are consistent with nucleotide changes occurring in SRA sequencing datasets (see **Figure 1D**) and with the APOBEC3G-mediating RNA editing mechanisms. To deduce aminoacid changes as consequences of these nucleotide changes, we analyzed codon changes occurring in the aggregated GISAID variants and we predicted its consequences by using the SnpEff program (30). Occurrences per variant type demonstrate that missense and synonymous variant occurrences are more frequent compared to frameshift/nonsense variant occurrences per genome (**Figure 2C**). Overall, missense/silent ratio of GISAID aggregated variants is 1.82, revealing a greater diversity in missense variants as well (**Supplementary Figure 1**). Consequently, codon change analysis demonstrates frequent non-neutral changes in the second position of the codons ACA>ATA, ACT>ATT and GCT>GTT and leads to missense variants Thr>Ile and Ala>Val, respectively. Also, non-neutral changes in the first position of codons CTT>TTT and GTT>TTT are frequent, leading to Leu>Phe and Val>Phe changes, respectively (**Figure 2D**). Almost all these changes can be explained by C>T (C>U) transition, already reported for SARS-CoV-2 protein changes. To understand these changes at the global population level, we calculated nucleotide diversity in nonsynonymous (πN) and synonymous (πS) sites of every SARS-CoV-2 protein across six different populations (Asia, Oceania, Europe, Africa, North America, and South America) by using GISAID genomes per geographical region.

**Figure 2:**
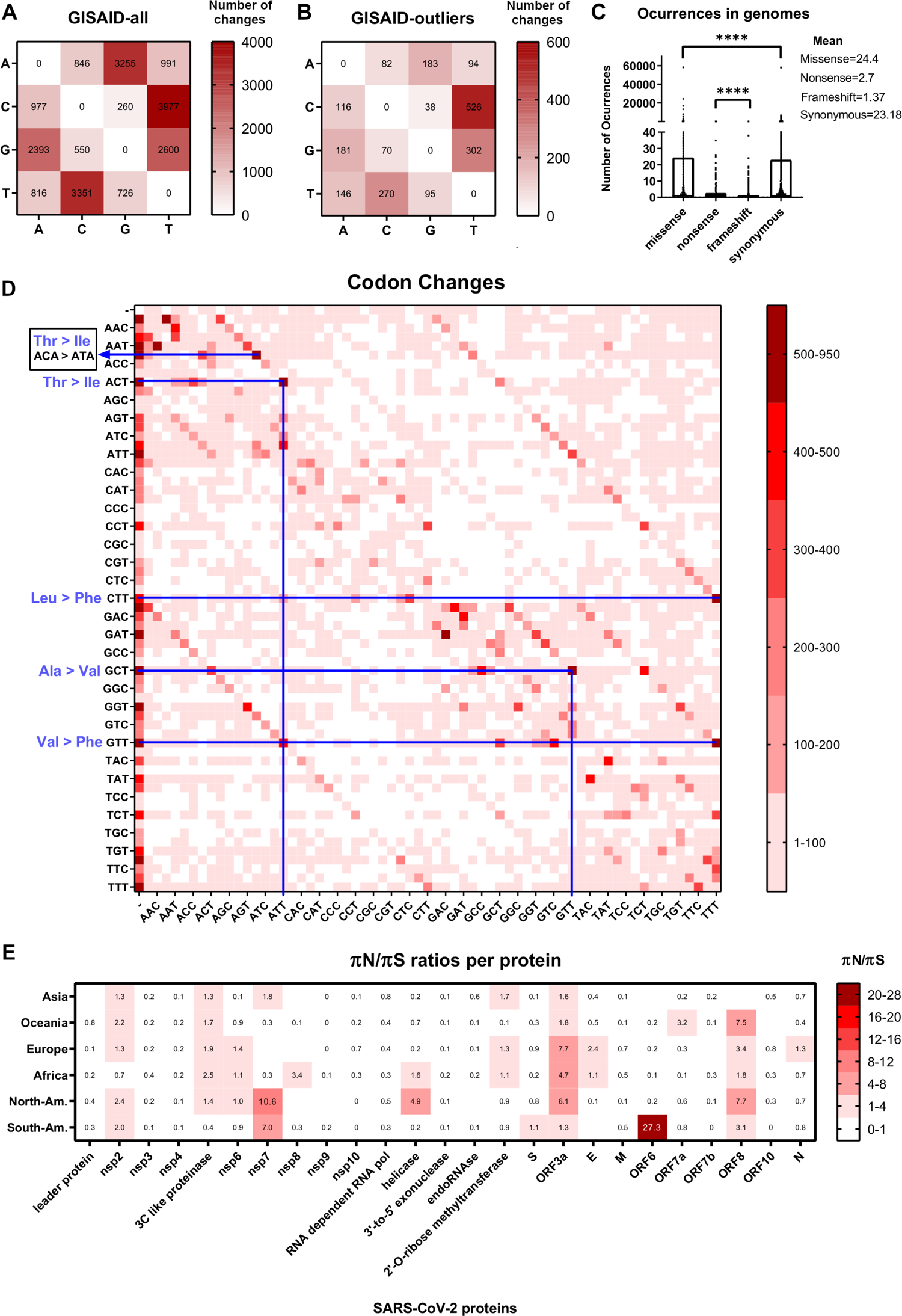
Non-neutral codon changes actively shape evolution of several SARS-CoV-2 proteins. A) Nucleotide change frequencies from 76,553 aggregated GISAD genome variants annotated with SnpEff program. Frequency boxes are colored from white to dark red as number of changes increases. **B**) Same as A for 243 aggregated GISAID genomes, corresponding to GISAID outlier samples. Frequencies boxes are colored from white to dark red, as the number of changes increases. **C**) Missense, nonsense, frameshift, and synonymous number of occurrences in 76553 GISAID genomes. Significance of comparisons were assessed with Mann-Whitney test (P<0.05 *, P<0.01 **, P<0.001 ***, P>0.05 ns). **D**) Plot of change frequencies across 32 codons in SARS-CoV-2. Changes are grouped in six categories and colored from light to dark red, according to the number of changes. **E**) Nonsynonymous/synonymous nucleotide diversity calculated by SNPGenie program for each SARS-CoV-2 protein across six geographical regions (Asia, Oceania, Europe, Africa, North America, and South America, respectively). Ratios are grouped in seven categories and colored from white to dark red, according to the ratio numbers.

A given protein is evolving by natural selection if the πN/ πS ratio is over one. Conversely, if πN/ πS ratio is less than one, a given protein is considered to be undergoing purifying selection, as previously described (66–68). An excess of shared nonsynonymous changes (implying positive natural selection) is present in non-structural proteins nsp2, nsp7, 3C-like proteinase, ORF3a and ORF8, respectively (**Figure 2E**). Conversely, purifying selection is present in the RNA-dependent RNA polymerase, Membrane protein (M), 3’-to 5’-exonuclease, non-structural proteins nsp3 and nsp4, respectively. Remarkably, ORF6 and to a lesser extent the Spike protein (S) of SARS-CoV-2 are actively evolving in South America.

### V1176F variant occurring in the Spike protein is predicted to improve fitness of Spike protein complex and is likely a novel SARS-CoV-2 mutation associated with increased mortality

We analyzed SARS-CoV-2 fixed alleles shared with at least a 3% variant frequency (VF) in one of the five referred to continental populations until August 3, 2020. The merge encompassed 51 variants. Consistent with intra-host variation, more than half of these changes lead to missense variants. Non-structural proteins nsp2, nsp3, nsp6 and nsp7 harbors Leu>Phe and Thr>Ile as frequent changes as seen in SARS-CoV-2 codon changes frequencies (**Figure 3A**). Also, consistent with the increased observed πN/ πS ratios per population, ORF3a and ORF8 display missense variants with high viral frequencies. Some variants are shared among populations with high viral frequencies, such as D614G variant in Spike protein, P323L variant in RNA-dependent RNA polymerase and RG203KR variant in Nucleocapsid. Of these, D614G variant in the Spike protein fulfill the category of mutation since this variant is positively associated with mortality and increases SARS-CoV-2 infectivity, as previously reported (18, 19, 69). As D614G was a rapidly emergent variant, this suggests novel emergent variants can also be mutations. Of importance, two more novel missense variants occurring in the Spike protein (S477N and V1176F) are exclusively from Oceania and South America (**Figure 3A**, red variants). To gain understanding of the effects of these variants, we calculated the Gibbs’ free energy associated with the variant changes (ΔΔG; ΔG_wild-type_ – ΔG_variant_) from all missense variants listed in Figure 3A using SARS-CoV-2 protein structures generated by the C-I-TASSER pipeline as inputs for the Foldx5 program (42–47). While most of the changes are predicted to be neutral or slightly stabilizing/destabilizing, there are more changes predicted to be energetically unfavorable rather than favorable, such as occurring in the RNA-polymerase (A97V), ORF3a (G251V) and Nucleocapsid (I292T). Conversely, other changes occurring in the non-structural proteins nsp3 (T1198K), nsp4 (F308Y) and Spike protein (D614G, V1176F) are predicted to stabilize these proteins (**Figure 3B**). The emergent V1176F variant, containing the recurrent signature Val>Phe (**Figure 2D**) is located at the stalk domain of Spike protein, specifically at the beginning of the heptad repeat 2 (HR2) domain (70). In agreement with a recent report, the D614G variant has a mildly stabilizing effect on protein stability but also alters protein dynamics according to mCSM-PPI2 analysis, since the predicted affinity change of the spike protein trimmer decreases (71). Conversely, the V1176F variant is predicted to increase affinity of the Spike protein trimmer (**Figure 3C, upper right**). Molecular dynamics simulations of the stalk domain trimmer demonstrate larger amplitude motions, since Root-mean-square deviations (RMSD) from wild-type stalk domain trimmer fluctuates in ∼1 nm (10 Å) over 20 nanoseconds of simulation. The V1176F variant increases this value after the same settings, increasing motility of the domain trimmer (∼1.4 nm, **Figure 3C middle right**). Also, the V1176F induces compactness of Stalk domain trimmer in ∼0.52 nm (5 Å) (**Figure 3C low right**) (72). Thus, the V1176F variant confers more flexibility to the Stalk trimmer domain. These observations agree with a recent report demonstrating extensive flexibility of this domain composed by three hinges in the pre-fusion model of the Spike protein, potentially necessary for enhanced binding mechanics with ligands such ACE2 (73). Of note, up until September 25, 2020 Brazil was the country with the highest frequency of genomes containing the V1176F variant, that was completely linked to the D614G mutation (**Figure 3D**). We also found four additional regions that present increasing cumulative distributions of genomes containing V1176F, including Scotland, USA, Australia, and Gibraltar that have occurred during global travel bans/restrictions (**Figure 3E**). Indeed, phylogenetic tree analysis containing genomes from Figure 3E demonstrate V1176F variant arose independently in clades G (containing S: D614G mutation), clade GH (containing ORF3a: Q57H variant) and clade GR (containing N: RG203KR variant) (**Figure 3F**). The predominant clade GR contain genomes from Brazil, Scotland, and Gibraltar while clade GH is represented exclusively of genomes from USA. Clade G is also present in Brazil and Scotland. Overall, this analysis suggests community transmission spread of V1176F occurred from independent sources rather than from a single source arising from travel. To correlate if this variant has an associated phenotype, we downloaded 7634 genomes with associated metadata from GISAID until September 28, 2020 and we grouped the genomes from released/deceased patients per country/region. Then, we performed Fisher’s exact test and identified variants with a significant difference in the viral frequencies between the groups (snpFreq program, available in the Galaxy server) (31, 35). Interestingly, a novel variant in the spike protein (QD613HG), leading to D614G mutation and variants occurring in the ORF3a (Q57H) and Nucleocapsid (RG203KR), are positively correlated with increased mortality ratios in Saudi-Arabia (**Figure 3H**). In Brazil, the D614G mutation along with the V1176F emergent variant in the Spike protein are also positively correlated with increased mortality ratios, including variants occurring in the nsp7, RNA-dependent RNA polymerase, and two emergent variants in South America occurring in the ORF6 and the nucleocapsid proteins (I33T and I292T variants, respectively, **Figure 3A**, **Figure 3G**). The RG203KR variants occurring in the Nucleocapsid was also found in Brazil. Four out seven of these variants occurring in Brazil (nsp7: L71F, S: V1176F, ORF6: I33T and N: I292T) are also positively correlated with increased mortality when comparing deceased patients versus Released + Hospitalized patients, suggesting these variants are robustly correlated with increased mortality in Brazil (**Figure 3H**). Of note, the V1176F variant arose independently and is not ligated to the I292T variant occurring in the Nucleocapsid (**Supplementary Table 4**). Thus, among emergent SARS-CoV-2 variants in Brazil, we prioritize the V1176F variant for further experimental study since it likely arose independently across SARS-CoV-2 clades in different countries, is predicted to improve fitness of the Spike protein and correlates with increased mortality ratios in Brazil.

**Figure 3:**
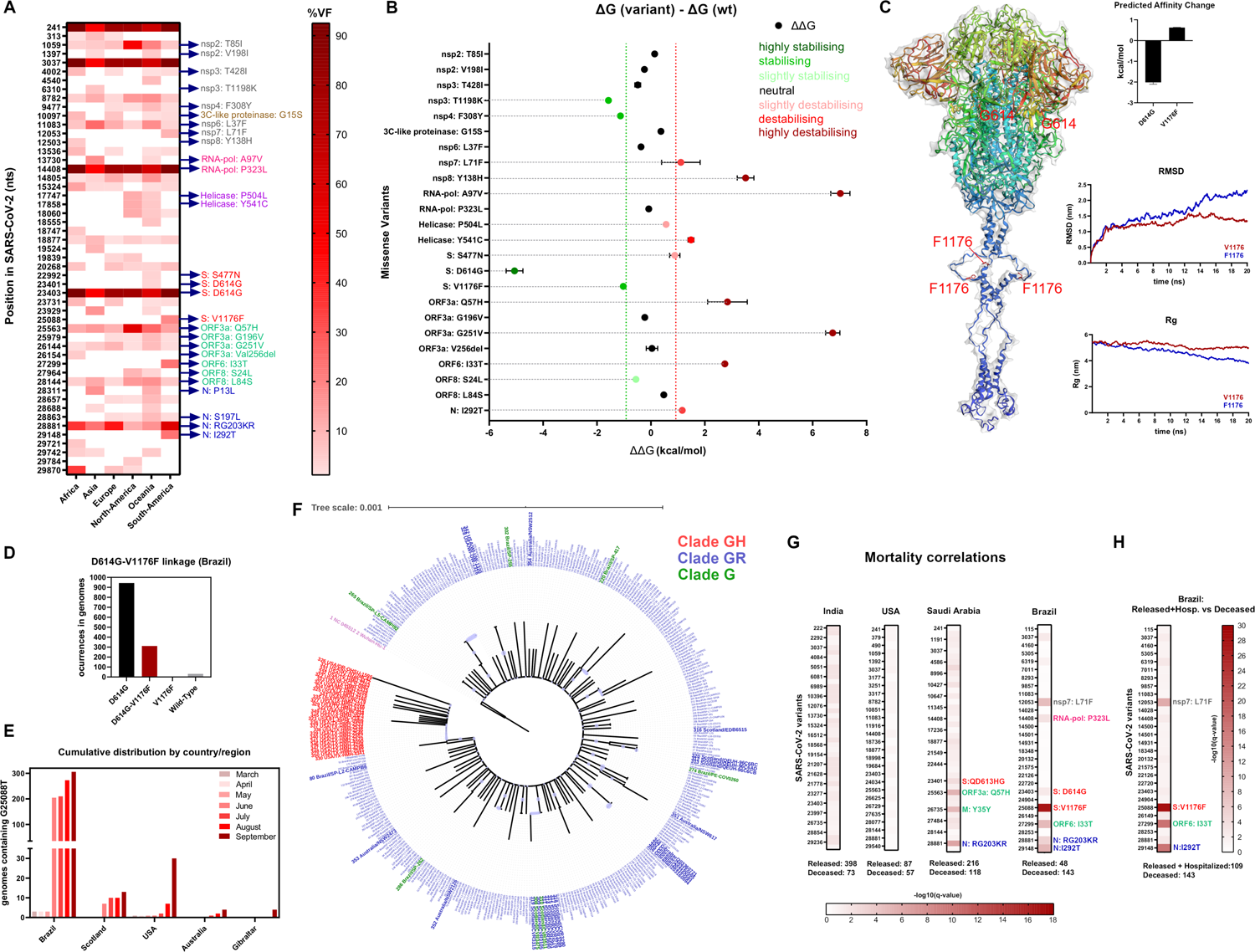
V1176F variant occurring in the Spike protein is predicted to improve fitness of Spike protein complex and is likely a novel SARS-CoV-2 mutation. **A**) Heatmap of viral frequencies from 51 variants shared at least 5% frequency within populations. Variants position in SARS-CoV-2 reference genome (Wuhan-Hu-1, GenBank Accession: MN908947.3) are depicted at left. At right, amino acid changes are colored by protein. **B**) Gibbs Free Energy Calculation (ΔΔG) of missense variants depicted in A, obtained by Foldx5 program. G (kcal/mol) was colored according to the energetic effect in SARS-CoV-2 structures depicted in the Y axis, as follows (in kcal/mol): highly stabilizing (< −1.84), stabilizing (−1.84 to −0.92), slightly stabilizing (−0.92 to −0.46), neutral (−0.46 to +0.46), slightly destabilizing (+0.46 to +0.92), destabilizing (+0.92 to +1.84), and highly destabilizing (> +1.84). **C**) (left) Rendered Spike protein trimmer structure with EzMol server using rainbow palette. D614G mutation and V1176F variant are depicted in red, including their sidechains. Volume is depicted in grey. The stalk domain of the Spike protein trimmer is depicted in dark blue. (Upper right) Predicted affinity change (kcal/mol) of Spike trimmer by mCSM-PPI2 server upon D614G and V1176F amino acid Spike protein changes. (middle right) RMSD values (in nanometers, nm) of 20 nanoseconds of simulation of the wild-type stalk domain trimmer (red) or the domain containing Phenylalanine in position 1176 (blue). (lower right) Radius of gyration values (in nanometers, nm) of the latter simulation. **D**) Occurrences per genome of the D614G mutation and V1176F variant alone or in combination in 974 Brazilian GISAID genomes until September 25, 2020. Complete linkage of V1176F variant with D614G mutation was found since non a single genome contains V1176F variant alone. **E**) Cumulative distributions of 358 genomes containing V1176F variant (G25088T) from January 2020 until September 28, 2020 in five geographical regions, respectively. **F**) Phylogenetic tree of the 358 genomes from E, constructed by Neighbor-Joining method, and visualized with the iTOL server. The genomes were colored by SARS-CoV-2 clades as follows: G (containing S: D614G mutation, in green), GH (containing ORF3a: Q57H variant, in red) and GR (containing N: RG203KR variant, in blue). Brazilian genomes belonging clade GR were depicted with small blue font and the other genomes were highlighted with higher font, for visualization purposes. **G**) Mortality correlations associated with SARS-CoV-2 variants in Released vs Deceased patients occurring in India, USA, Saudi Arabia, and Brazil until September 25, 2020, respectively. The corrected p-values from fisher exact test (q-values) were obtained from the snpFreq program, available in the Galaxy server and plotted as the negative logarithm in base 10 of each q-value (significance: q-value>0.005). Significant variants were depicted at the right of each bar. **H**) Same as G, but with the comparison Released + Hospitalized vs Deceased patients in Brazil.

### Variant S447N occurring in the Spike protein is a novel mutation that increases Spike-ACE2 binding and is associated with higher worldwide fatality rates

We next addressed whether the referred variants are associated with higher mortality ratios in Brazil and Saudi-Arabia, as well as other emergent variants in the Spike protein, are correlated with higher fatality ratios worldwide until September 28, 2020. Among the refereed variants, the I292T variant occurring in the nucleocapsid is associated with higher fatality rates across several countries (p<0.033, Spearman correlation, **Figure 4A**, **Supplementary Table 5**). Eleven variants occurring in the Spike protein were present in more than four countries and out of them, variants A222V, S477N and E780Q are positively correlated with increased fatality ratios. These findings are consistent with previous reports for the D614G mutation that is also positively correlated with higher fatality ratios (**Figure 4B**) (19). The V1176F variant was not found to be correlated with increased fatality ratios across the world, probably due to highly unbiased viral frequencies across the world. Up until August 3, 2020, the S477N spike variant that emerged in Oceania (Australia) had a low viral population frequency (∼ 3.5%, **Figure 3A**) along with the A222V variant being absent or maintained at a very low frequency. However, a dramatic increase of cumulative genomes was observed between the months June and July 2020 for both variants, not seen for the I292T variant occurring in the nucleocapsid (**Figure 4C**). The rapid emergence of the S477N and A222V variants correlates with the recent second wave of COVID19 that has occurred in Australia and the United Kingdom (**Figure 4D**, left and right, respectively). Nevertheless, this rapid increase in the population frequencies of these variants could be due to the founder effect usually seen in outbreaks, and not due increased fitness of SARS-CoV-2 (see countries in Figure 4B, A222V and S477N, respectively) (69). To discard the former founder effect, we examined GISAID clade frequency of both variants, demonstrating variant A222V arose independently in all major SARS-CoV-2 clades (74) and variant S477N arose in clades G, GH and GR, respectively (**Figure 4E**, upper and lower, respectively). Variant A222V occurred in the N-terminal domain of the Spike protein (light brown domain, **Figure 4F**, left) and variant S477N occur in the Receptor Binding Domain (RBD) of the Spike protein (purple domain, **Figure 4F**, left). This variant is located near the interface between ACE2 and the RBD, the latter is expected to cause enhanced binding of the RBD to the ACE2 human receptor (**Figure 4F**, right). We replaced serine for asparagine in position 477 in the RBD (complexed with ACE2 dimer) with foldx (46), and we calculated the predicted binding energy upon this change by using mCSM-PPI2 server (49). The change is predicted to add one more polar interaction when asparagine is present in the RBD, increasing the affinity between RBD and ACE2 (**Figure 4G**). We examined the deep mutational scanning of amino acid changes in the RBD performed by Starr et al (75) in a high throughput yeast-surface-display system for measuring expression of folded RBD protein and its binding to ACE2 (76). Among fourteen variants occurring in the RBD, only variant S477N increased both expression of the RBD, a parameter positively correlated with folding (**Figure 4H**) and its binding to ACE2, respectively (**Figure 4I**). The combination of these two properties can lead to the generation of a more infectious viruse, explaining to a large extent the dramatic increase of the S477N variant in Australia.

**Figure 4:**
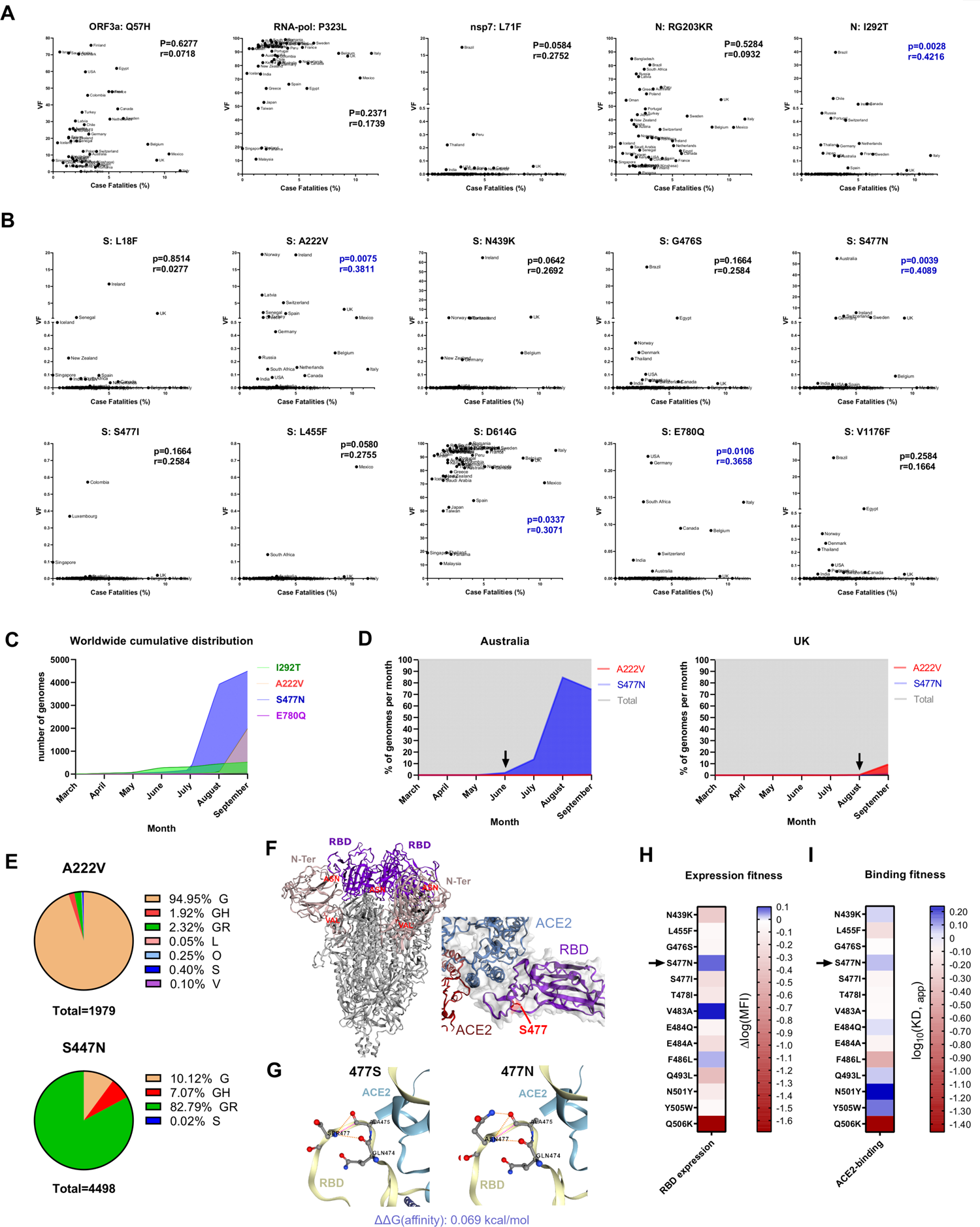
Variant S447N occurring in the Spike protein is a novel mutation that increase Spike-ACE2 binding and is associated with worldwide higher fatality rates. A) Correlation analysis of COVID-19 fatality rates among 49 countries against variant frequencies occurring in various SARS-CoV-2 proteins: ORF3a (Q57H), RNA-dependent RNA polymerase (P323L), nsp7 (L71F), N (RG203KR) and N (I292T), respectively. Spearman’s correlation coefficients (r) were calculated, including confident p-values (P) to discard random sampling. The significance thresholds were the following: P<0.05 *, P<0.01 **, P<0.001 ***, P<0.0001 ****, P>0.05 ns. China were discarded from these analyses due its large population size and consistently low number of COVID19 cases since early April 2020. Fatality rate data was obtained from John Hopkins coronavirus resource center (https://coronavirus.jhu.edu/map.html), accessed on September 28, 2020. **B**) Same analysis as A, for five emergent variants occurring in the Receptor Binding Domain (RBD) portion of the Spike protein (N439K-L455F), including variants L18F, A222V, D614G, E780Q and V1176F occurring in the Spike protein. **C**) Cumulative distributions over time of genomes containing variants I292T in the nucleocapsid and emergent variants A222V, S477N and E780Q in the spike protein, respectively. Cumulated genome numbers were plotted at the end of the indicated months. **D**) Frequency plots over time of cumulated genomes containing spike protein emerging variants A222V, S477N occurring in Australia (left) and United Kingdom (right), respectively. Cumulated frequencies were plotted at the end of the indicated months. **E**) Graph chart indicating the clade frequencies of variants A222V (Upper) and S477N (lower), reported from GIDAID database, until September 28, 2020. **F**) (Left) Spike protein Trimmer indicating positions of variants N477 and V222 (red letters). Variant N477 locates in the Receptor Binding Domain (RBD, residues 331-530, depicted in purple) and Variant V222 locates in the N-terminal domain (residues 1-316, depicted in light brown). The Spike structure was shown as cartoon with grey color, obtained with EzMol server. (Right) Magnification of the interaction between the RBD (purple) with ACE2 dimer (blue and red structures, respectively) showing the contact surfaces. Variant N477 and its surface is depicted in red. The image was rendered in the EzMol server. **G**) Polar interactions of S477 (left, wild type) and N477 (right, mutant) depicted by discontinued lines in the RBD. The polar amino acid interactions of S477 and N477, including the predicted ΔΔG (affinity, kcal/mol) between the wild-type or mutant RBD with ACE2 dimer were obtained by using the mCSM-PPI2 server. Positive values of ΔΔG indicate increasing affinity between the RBD and ACE2 dimer. **H**) Expression fitness of fourteen emergent variants occurring in the RBD domain of the spike protein (see Supplementary Figure 2), measured by high throughput yeast-surface-display system. Expression measurements were plotted as the difference in log-mean fluorescence intensity (MFI) relative to wild-type (logMFI =Δ logMFI_variant_ -logMFI_wild-type_). Positive values (blue) indicate higher RBD expression, therefore higher folding fitness. Expression data was obtained from here: https://jbloomlab.github.io/SARS-CoV-2-RBD_DMS/. I) ACE2 binding fitness of the referred RBD variants, measured by high throughput yeast-surface-display system. Binding measurements were plotted as the difference in log10(KD, apparent) relative to wild-type (log10(KD, apparent) = log10(KD, apparent)_wild-type_ – log10(KD,app)_variant_). Positive values (blue) indicate higher affinity between RBD and ACE2. Binding data was obtained from here: https://jbloomlab.github.io/SARS-CoV-2-RBD_DMS/.

In conclusion, we have obtained evidence that the S477N variant is a gain of function mutation occurring in the Spike protein, that is positively correlated with increased fatality rates and is becoming dominant as its increases, as was also recently observed for the D614G mutation that became dominant throughout the world (18).

## Discussion

In this study we aimed to analyze over 94,000 SARS-CoV-2 sequences deposited between GISAID and SRA databases within the first eight months of this pandemic (up until the beginning of August 2020). We characterized the existence of intra-host viral hypermutation that results in an excessive number of variants per genome in less than 2% of SARS-CoV-2 sequences (**Figure 1A** and **1B**, respectively). This phenomenon was already described for HIV-1 virus *in vivo*, demonstrating that HIV-1 reverse transcriptase contributed only to 2% of mutations, and the majority was caused by host cytidine deaminases of the A3 family mediated editing (77). Here, we present evidence that enzymatic RNA editing in combination with microdiversity contributes to SARS-CoV-2 diversity at a global level, leading to more than 23,000 major viral frequency variants within 76,000 GISAID genomes. In SARS-CoV-2 genomes, the C>T (C>U) transversion is substantially present both in hypermutants and non-hypermutant samples, suggesting APOBEC3G mediated RNA editing involvement, as previously reported in smaller sample sizes (59, 60). Also, the A>G transversion is also present overall but not in hypermutant genomes (Figures **1D**, **2A** and **2B**, respectively), implying an active role of ADAR deaminases during SARS-CoV-2 infection (78). We argue ADAR-mediating RNA editing is not the main enzyme involved in the hypermutation mechanism, but rather APOBEC3G deaminase complexes. Also, we have observed substantial G>T transversions in SARS-CoV-2 genomes. This transversion has been already reported for other RNA viruses such as Maize streak virus (79) and is has been linked with the formation of 8-oxoguanine, known to be the most common cause of spontaneous G>T (G>U) transversions in RNA (80). Recently, it has been reported that tissue damage from neutrophils induces oxidative stress upon SARS-CoV-2 infection (81), implying that reactive oxidative species (ROS) mediated mutagenesis is likely the mechanism that cause this transversion. The hypermutated SARS-CoV-2 variant signature often contains nonsense variants that are predicted to inactivate several SARS-CoV-2 proteins, probably leading to an efficient mechanism of lethal mutagenesis to control viral spread (**Figure 1E**). In addition, other signatures are present at the intra-host level, implying microdiversity as another potential source of this variation (**Figure 2A** and **2B**, respectively). It is possible that these combined forces produce quasi-species of viruses with enough sequence diversity that may influence viral pathogenesis and drive SARS-CoV-2 evolution, sometimes also leading to viral extinction (82, 83). Overall, we propose that human host are major drivers of SARS-CoV-2 diversity rather than the virus itself, evidenced by the levels of intra-host variation and hypermutation at different degrees, both fueled by enzymatic RNA-editing mechanisms. We support the latter with the observed non-random signatures of nucleotide changes in these mechanisms, and the presence of SARS-CoV-2 error-correction machinery, not seen in other RNA viruses. Although we found a significant amount of intra-host variation in SARS-CoV-2, neutral evolutionary theory predicts most of these variants as having no or neutral effects (84). Nevertheless, positive, or negative selection can occur within-populations on viral variants, by conferring advantageous properties to viruses that ultimate lead to mutations. Here we have demonstrated positive natural selection of several SARS-CoV-2 proteins per population, occurring in nsp2, nsp7, 3C-like proteinase, ORF3a and ORF8 proteins and we highlighted mutations in the Spike protein, evolving in South America (Brazil, V1176F) and Oceania (Australia, S477N). To begin to understand how these variations may affect their encoded proteins, we assessed their structural consequences in protein models that demonstrate that these variants tend to cause more unfavorable than favorable energetic changes. This phenomenon has been observed in the spectrum of variants occurring in the RBD of the Spike protein, proving that most of the variants occurring in the RBD constraints its folding and binding to ACE2 (75). In the case of SARS-CoV-2, immunological pressure from host could lead to this type of phenomena as well and is consistent with the observed number of intra-host missense over synonymous variants, respectively. Conversely, we found missense variant V1176F, occurring in the Spike protein, is predicted to be energetically favorable and confers flexibility to the Stalk domain of the viral Spike protein trimmer, previously described to be important for Spike protein flexibility and binding to ACE2 (73). Phylogenetic analysis demonstrated V1176F variant likely emerged in South America and arose independently in several countries associated with the D614G mutation, suggesting that this variant is being positively selected among others occurring in the Spike protein (**Figure 3F**). This variant is also correlated with higher mortality ratios (**Figure 3G** and **3H**, respectively) and it is possible that it increases the fitness of SARS-CoV-2 infection by conferring flexibility to the stalk domain of the spike protein. The same conclusions applied to variant S477N, occurring in the RBD of the Spike protein: is energetically favorable in RBD-ACE2 binding and favor expression of the RBD, with the latter conclusions being experimentally supported. This mutation also arose independently in a noticeably short period of time and become dominant in Australia within two months (**Figure 4D**). In addition, the S477N variant is constantly spreading across European countries (**Figure 4B**) and correlates with higher mortality ratios. These observations provide strong evidence that the S477N variant is a novel gain of function Spike protein mutation, as has recently been demonstrated for the D614G mutation (18). We argue that the constant spread of V1176F and S477N variants over the world ultimately may lead to a further significant concern in public health, due to their association with higher mortality rates.

A remaining question is the association of higher fatality rates of the A222V variant occurring in the N-terminal domain of the Spike protein, and the I292T variant occurring in the Nucleocapsid, among others. It is possible that these variants can confer antigenic escape, since recently, it has been registered that a reinfection case containing A222V and D614G mutations has occurred (85). Nucleocapsid variation has also been documented in the nucleoprotein of the influenza virus (86) (87) and nucleocapsid of the hepatitis virus (88). Both RNA viruses escape cellular immunity by these mechanisms and could also be the case for SARS-CoV-2.

In summary, we have presented potential molecular mechanisms that help researchers to understand variation diversity fueled natural selection in SARS-CoV-2. It is important to continue to track emergent viral variants with the bioinformatics tools developed and highlighted in this manuscript since the evidence presented here lead us to propose V1776F and S477N variants are novel SARS-CoV-2 mutations, due to their positive correlations with increased fatality ratios, as previously evidenced with D614G mutation occurring in the Spike protein. Further conclusions concerning the effects of these variants on viral fitness and host mortality will be accomplished by future structure-function based studies using viral Spike protein mutants and studying effects on viral entry and *in vivo* rodent models expressing the human ACE2 receptor.

## Data Availability

Data and Code Availability 76,553 FASTA genomes and associated sequencing metadata were downloaded from GISAID database from January 1, 2019 until August 3, 2020, specifying human as source host (https://www.gisaid.org/). The associated sequencing metadata including major variants per sample are available at Supplementary Table 1. 974 Brazilian FASTA sequences were downloaded from GISAID database from January 1, 2019 until September 25, 2020, specifying human as source host and South America / Brazil as location. Acknowledgements to all laboratories/consortia involved in the generation of GISAID genomes used in this study are listed in Supplementary Table 2.17,560 sequencing datasets were downloaded from Sequence Read Archive Repository (SRA, https://www.ncbi.nlm.nih.gov/sars-cov-2/) From December 1, 2019 until July 28, 2020. Associated sequencing run accessions, sequencing metadata and related BioProjects are listed in Supplementary Table 3. The code generated during this study to replicate most of the computational calculations performed in this manuscript is available at the following github repository: https://github.com/cfarkas/SARS-CoV-2-freebayes.

## Acknowledgments

Powered@NLHPC: This research was partially supported by the supercomputing infrastructure of the NLHPC (ECM-02). This research was partially funded by research funding from the CIHR, Research Manitoba and the CancerCare MB Research Foundation.

## Author Contributions

CF conceived of this study and performed all bioinformatics analysis and wrote the manuscript. AM performed mutant Spike protein analysis and assist in biophysical studies. JH assisted study design, data interpretation and manuscript writing.

## Supporting information

**Supplementary Table 1:** Sequencing metadata of 76554 GISAID genomes downloaded until August 3, 2020. For every GISAID genome, we provided GISAID genome name, GISAID unique identifier (Accession ID), geographic location, host, sequencing technology, lineage, and clade fields, among other information. The last column indicates the number of variants per genome (Major viral variants).

**Supplementary Table 2:** Acknowledgements from sequencing laboratories and/or consortia associated with GISAID genomes listed in Supplementary Table 1, plus genomes downloaded until September 28, 2020, containing variants V1176F, S477N and A222V occurring in the Spike protein.

**Supplementary Table 3:** Sequencing metadata of 17560 Sequencing Read Archive (SRA) datasets downloaded until July 28, 2020. For every SRA dataset, we provided NCBI run accession, Assay type (indicates if amplicon, RNA-seq u other sequencing corresponds), sequencing size (bases, in nucleotides), Biosample accession ID, Center Name (depositor), release date, SRA study accession, BioProject and geographic location, among other information. The last column indicates the number of variants per sample (Major viral variants, viral frequency > 0.5).

**Supplementary Table 4:** Sequencing metadata of 543 and 393 GISAID genomes containing variants N: I292T and S: V1176F, respectively. We accessed GISAID database on September 28, 2020 and we downloaded genomes containing the variant I292T or V1176F. The associated metadata from both cohorts are presented in this table. For every GISAID genome, we provided GISAID genome name, GISAID unique identifier (Accession ID), collection information, geographic location, host, sequencing technology, lineage, and clade, among other information. We used GISAID unique identifiers to overlap both groups. No overlap was found.

**Supplementary Table 5:** Worldwide fatality ratios per country, related to Supplementary Figure 2. Worldwide fatality ratios among 49 countries obtained from John Hopkins coronavirus resource center (https://coronavirus.jhu.edu/map.html), accessed on September 28, 2020. At right, we calculated viral allele frequencies of several SARS-CoV-2 variants per country, based on the GISAID database, also accessed on September 28, 2020. We analyzed the following variants: ORF3a (Q57H), N (RG203KR), RNA-dependent RNA polymerase (P323L), S (D614G), S (V1176F), nsp7 (L71F) and N (I292T), respectively.

**Supplementary Figure 1:** Intra Host variant effects. 23,269 aggregated variants from 76554 GISAID genomes were merged as indicated in Figure 2A. These variants were classified by SnpEff program, available in the Galaxy server. We plotted a heatmap with the number of changes per consequence type (see x-axis) against every SARS-CoV-2 protein (see y-axis). We also calculated the overall missense/silent ratio occurring in SARS-CoV-2 proteins (1.82).

