## Supplementary Figure 1 for "Large-scale population analysis of SARS-CoV-2 whole genome sequences reveals host-mediated viral evolution with emergence of mutations in the viral Spike protein associated with elevated mortality rates"

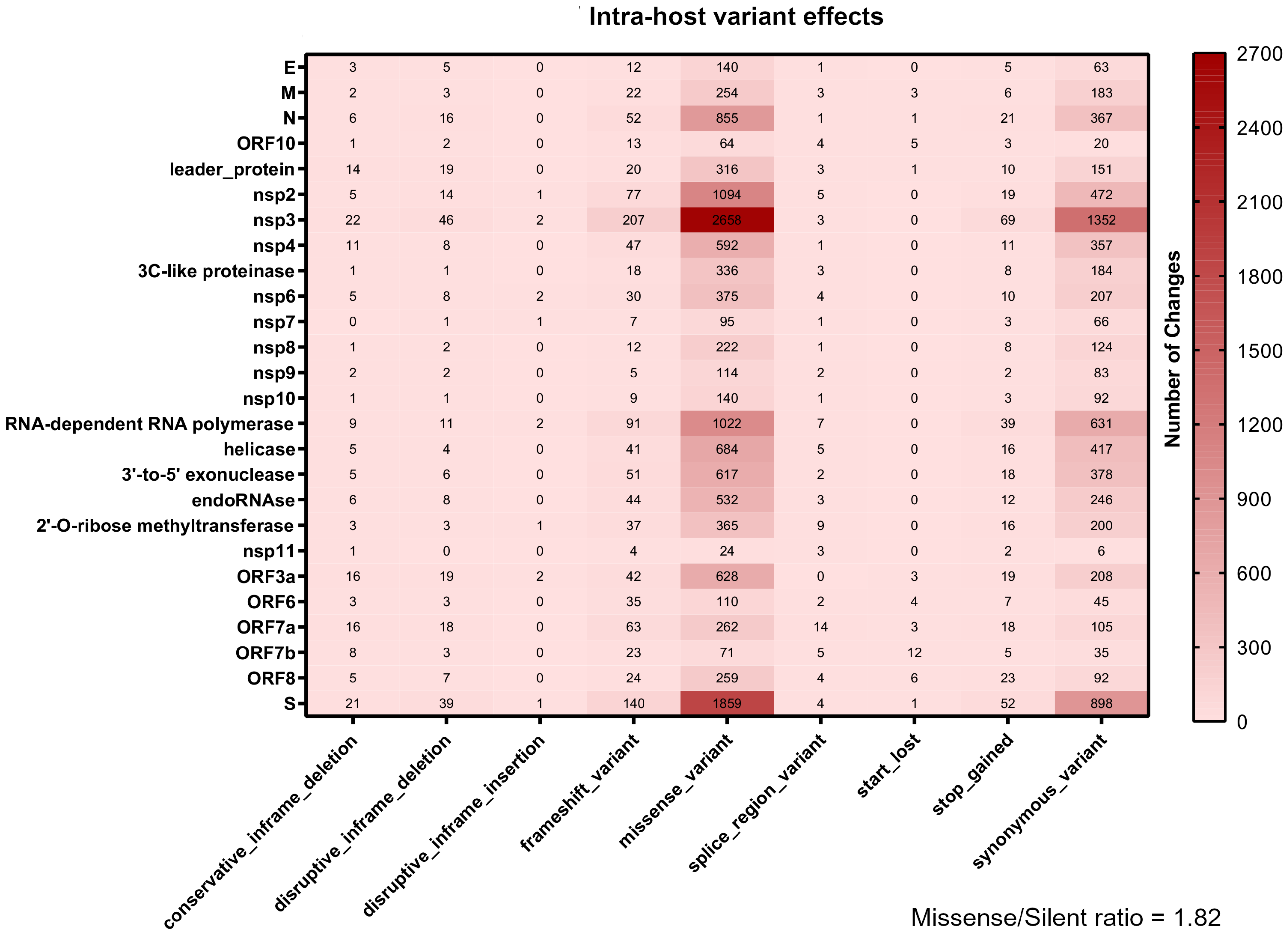


| | conservative_inframe_deletion | disruptive_inframe_deletion | disruptive_inframe_insertion | frameshift_variant | missense_variant | splice_region_variant | start_lost | stop_gained | synonymous_variant |
|---|---|---|---|---|---|---|---|---|---|
| E | 3 | 5 | 0 | 12 | 140 | 1 | 0 | 5 | 63 |
| M | 2 | 3 | 0 | 22 | 254 | 3 | 3 | 6 | 183 |
| N | 6 | 16 | 0 | 52 | 855 | 1 | 1 | 21 | 367 |
| ORF10 | 1 | 2 | 0 | 13 | 64 | 4 | 5 | 3 | 20 |
| leader_protein | 14 | 19 | 0 | 20 | 316 | 3 | 1 | 10 | 151 |
| nsp2 | 5 | 14 | 1 | 77 | 1094 | 5 | 0 | 19 | 472 |
| nsp3 | 22 | 46 | 2 | 207 | 2658 | 3 | 0 | 69 | 1352 |
| nsp4 | 11 | 8 | 0 | 47 | 592 | 1 | 0 | 11 | 357 |
| 3C-like proteinase | 1 | 1 | 0 | 18 | 336 | 3 | 0 | 8 | 184 |
| nsp6 | 5 | 8 | 2 | 30 | 375 | 4 | 0 | 10 | 207 |
| nsp7 | 0 | 1 | 1 | 7 | 95 | 1 | 0 | 3 | 66 |
| nsp8 | 1 | 2 | 0 | 12 | 222 | 1 | 0 | 8 | 124 |
| nsp9 | 2 | 2 | 0 | 5 | 114 | 2 | 0 | 2 | 83 |
| nsp10 | 1 | 1 | 0 | 9 | 140 | 1 | 0 | 3 | 92 |
| RNA-dependent RNA polymerase | 9 | 11 | 2 | 91 | 1022 | 7 | 0 | 39 | 631 |
| helicase | 5 | 4 | 0 | 41 | 684 | 5 | 0 | 16 | 417 |
| 3'-to-5' exonuclease | 5 | 6 | 0 | 51 | 617 | 2 | 0 | 18 | 378 |
| endoRNAse | 6 | 8 | 0 | 44 | 532 | 3 | 0 | 12 | 246 |
| 2'-O-ribose methyltransferase | 3 | 3 | 1 | 37 | 365 | 9 | 0 | 16 | 200 |
| nsp11 | 1 | 0 | 0 | 4 | 24 | 3 | 0 | 2 | 6 |
| ORF3a | 16 | 19 | 2 | 42 | 628 | 0 | 3 | 19 | 208 |
| ORF6 | 3 | 3 | 0 | 35 | 110 | 2 | 4 | 7 | 45 |
| ORF7a | 16 | 18 | 0 | 63 | 262 | 14 | 3 | 18 | 105 |
| ORF7b | 8 | 3 | 0 | 23 | 71 | 5 | 12 | 5 | 35 |
| ORF8 | 5 | 7 | 0 | 24 | 259 | 4 | 6 | 23 | 92 |
| S | 21 | 39 | 1 | 140 | 1859 | 4 | 1 | 52 | 898 |
